# Breath-borne VOC Biomarkers for COVID-19

**DOI:** 10.1101/2020.06.21.20136523

**Authors:** Haoxuan Chen, Xiao Qi, Jianxin Ma, Chunyang Zhang, Huasong Feng, Maosheng Yao

## Abstract

Rapid diagnosis of COVID-19 is key to controlling the pandemic. Here we report the potential breath-borne volatile organic compound (VOC) biomarkers for COVID-19. Higher levels of ethyl butanoate were detected in exhaled breath of COVID-19 patients (N=10) than healthy controls/health care workers (N=21), lung cancer (LC) patients (N=7) and backgrounds. In contrast, breath-borne butyraldehyde and isopropanol (an efficient SARS-CoV-2 inactivation agent) were significantly higher for non-COVID-19 respiratory infections (URTI) (N=22) than COVID-19, HC, LC patients and backgrounds. Breath-borne isopropanol emission from COVID-19 patients varied greatly up to ∼100-fold difference. COVID-19 patients had lower acetone levels than other subjects, except LC patients. The monitoring of ethyl butanoate, butyraldehyde and isopropanol could lend considerable support in rapidly screening COVID-19; and alerting the presence of COVID-19 patient in particular environments.

**One Sentence Summary:** COVID-19 patients emit distinctive VOC profiles

## Main Text

The COVID-19 pandemic has already caused significant loss of life, social disruption and economic standstill. The crisis is still continuing at an alarming speed throughout much of the world. As of now, the future of this pandemic remains to be largely uncertain. Diagnosis and screening of the COVID-19 is central to stemming the pandemic. Presently, reverse transcription-polymerase chain reaction (RT-PCR) is mainly used together with throat and/or nasal swabs for diagnosis and hospital discharges of COVID-19.^1^ However, the technique requires lengthy procedures and many bio-agents; and it often unfortunately leads to false negatives.^1^ A computed tomography scan (CT) is sometimes used to supplement RT-PCR.^2,3^ For asymptomatic patients yet posing a hidden infection risk, the accuracy of current methods need to be investigated and validated urgently.^1^ In clinical settings, it is challenging to render a decision for discharging a COVID-19 patient solely based on the results from throat or nasal swabs.^4^ It is equally important, yet no methods available, to monitor the public environments for its SARS-CoV-2 safety. The world is now historically at the cross road of opening up economies while tolerating the risk of another wave of the COVID-19 spread. This work is conduced to search for a specific biomarker for COVID-19 that can be used non-invasively to rapidly screen COVID-19 patients even before they develop symptoms. We recruited a total of 60 subjects (Tables S1-S5) as described in details in Methods. Breath-borne volatile organic compounds (VOCs) (N=40 species) profiles from these 60 subjects and related backgrounds (N=30) were analyzed using a commercial breath VOC analyzer.

Here we for the first time report the potential breath-borne VOC biomarkers for COVID-19. The COVID-19 patients were shown to have statistically significant higher levels of ethyl butyrate (29.13, 95.67) (95% CI, N=10) than heathy control (16.00, 24.3) (95% CI, N=12) (p-value=0.019), HCW (12.07, 31.05) (95% CI, N=9) (p-value=0.041), and LC (−37.65, 12.23) (95% CI, N=7), except URTI patients (24.77, 63.87) (95% CI, N=22) (p-value=0.341) (Table 1, Table S7, Fig 1A, B). There was no statistically significant difference detected in ethyl butyrate between background(ward) air and exhaled breath from LC patients (p-value=0.356). In addition, COVID-19 patients had statistically significant lower levels of isopropanol-1 (RI: 920.7; Dt: 1.2224) than healthy controls (p-value=0.0329) and URTI patients (p-value<0.001), but not HCW (p-value=0.838) and LC patients (p-value=0.238) (Table S6, Table 1). Examples of VOC profiles were presented for three groups in Fig 1B. The peaks observed for both propanol and ethyl butyrate were further confirmed using their standards (more than 99% purity grade) (Fig 1B) as stated in Methods. In contrast, acetone levels for COVID-19 patients were observed to be substantially lower than those of HC, HCW, and URTI patients (p-values<0.016; Table 1, Fig. 1C). For ethyl butyrate, all subjects except LC patients, had statistically significant higher levels than the backgrounds (p-values<0.0051; Table 1). No statistically significant differences were observed for propanol-1 and propanol-2 (numbers refer to different isomers) between COVID-19 patients and healthy controls (p-values>0.277; Table 1, Fig S1). It seems lower acetone and higher ethyl butyrate levels were distinctive for COVID-19 patients.

**Table 1.**
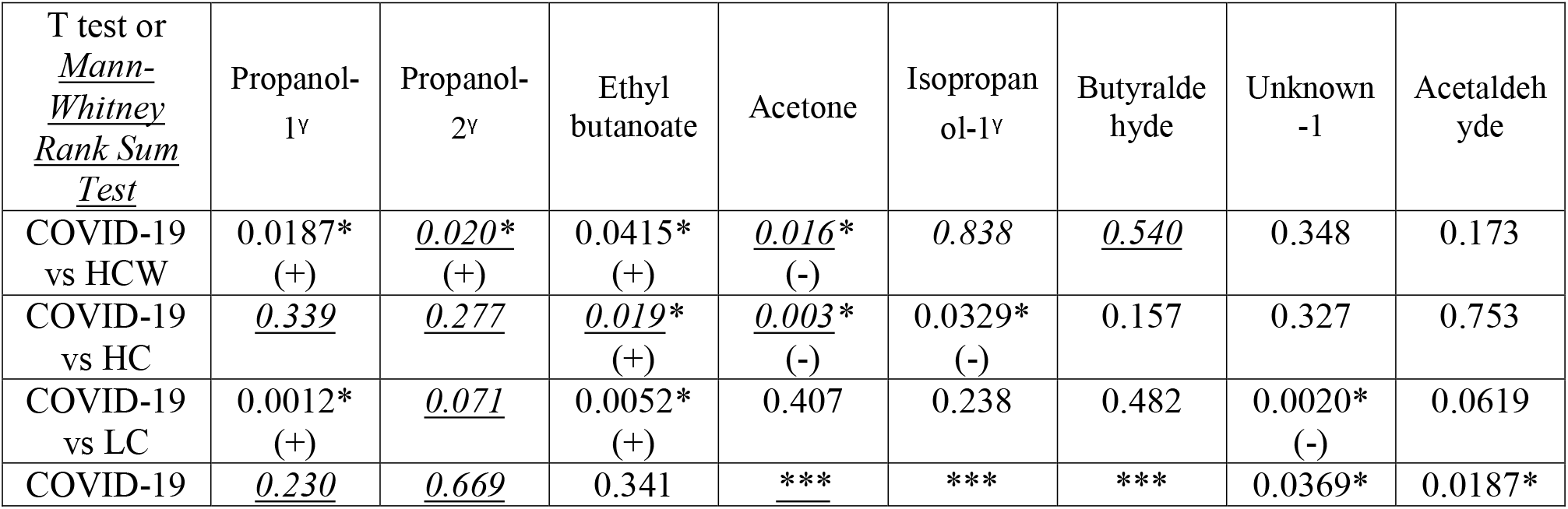
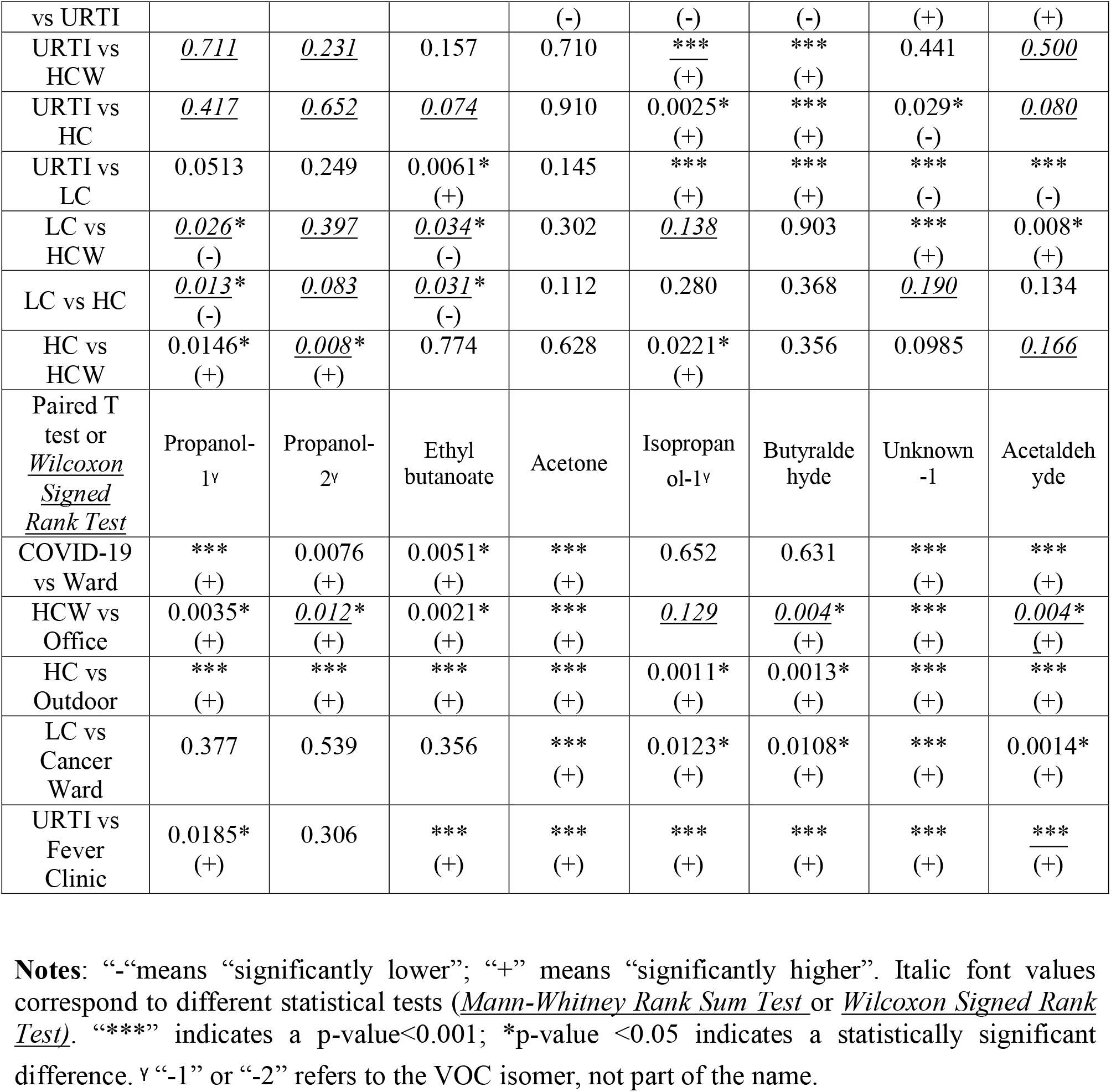
Summaries (p-values) of unique breath-borne VOC species profiles detected using gas chromatography-ion mobility spectrometry (GC-IMS) for COVID-19, healthy controls, HCW, URTI, LC patients, and backgrounds (Tables S1-S7).

**Fig 1.**
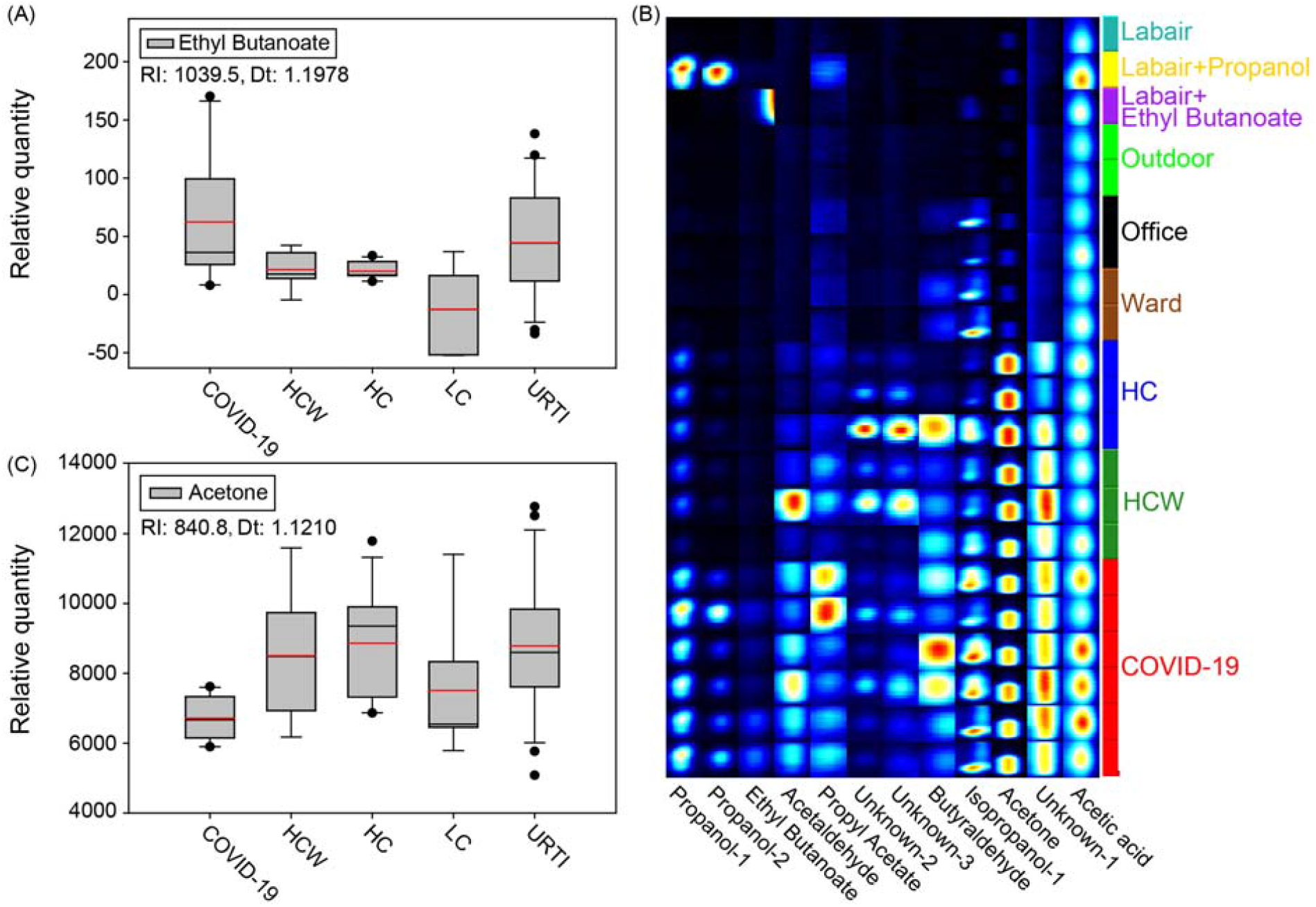
Characteristic breath-borne VOC boxplot profiles for COVID-19 patients compared to other groups of subjects: A) Ethyl butyrate levels of COVID-19 patients compared to those in other four groups; B) Acetone levels of COVID-19 patients compared to those of other four groups; C) Examples of VOC peak prints for selected COVID-19 patients, controls and various environments. For unknown VOCs, their Retention Index (RI) and Drift Time (Dt) values are listed in Table S7. “Labair+propanol/ethyl butanoate” indicates the standard spikes into the background lab air.

In contrast with COVID-19 patients, isopropanol-1 and butyraldehyde levels were detected to have substantially increased for URTI patients (N=22) (Fig 2; Table 1). In averages, the URTI patients had the highest isopropanol-1 and butyraldehyde levels, followed by HC, LC, HCW, and COVID-19 patients (Fig 2; Table 1). Examples of VOC abundance prints are shown for URTI patients in Fig 2B. Additionally, it seems that both isopropanol-1 and butyraldehyde followed similar patterns with respect to these five groups of subjects (Fig 2 A, C). For butyraldehyde, no statistically significant differences were detected between other subjects other than URTI groups. Breath-borne isopropanol-1 and butyraldehyde for UTRI patients were significantly higher than those measured from the backgrounds (p-values<0.001; Table 1). The peak of ethyl butyrate in Fig 2B was due to its calibration performed right before isopropanol. Lung cancer patients, clearly different from other groups of subjects, were shown to generally have higher average levels of Unknown-1 (RI:741.3; Dt: 1.0657) and acetaldehyde (Fig 3). It seems that no specific candidate VOCs were statistically different between LC patients and healthy controls (Fig 3; Table 1).

**Fig 2.**
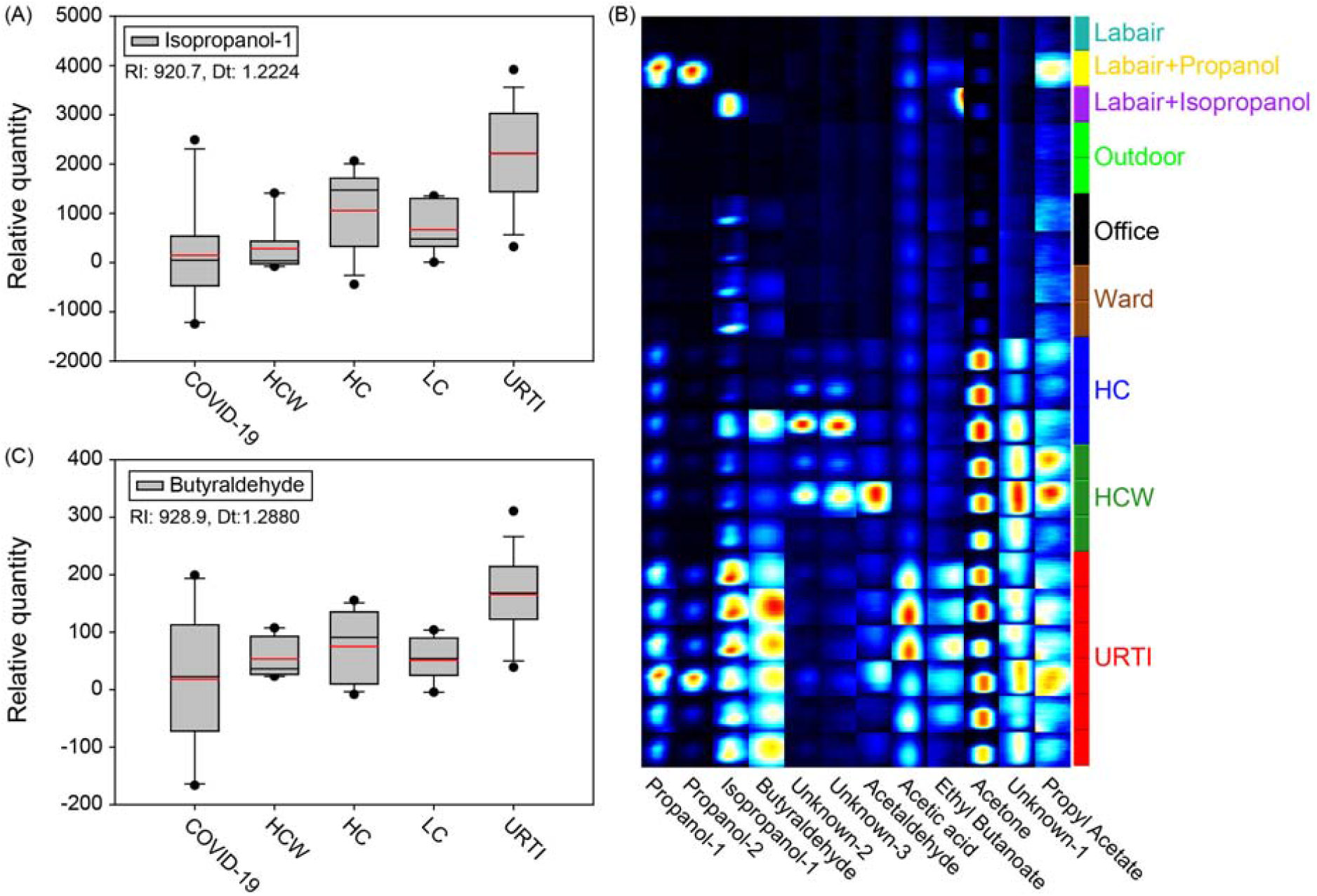
Characteristic breath-borne VOC boxplot profiles for the URTI (non-COVID-19) patients compared to other groups of subjects: A) Isopropanol-1 levels of URTI patients compared to those in other different groups; B) Butyraldehyde levels of URTI patients compared to those in other groups; C) Examples of VOC abundance prints for selected URTI patients, controls, various environments, isopropanol-1 and propanol standards. For unknown VOCs, their RI and Drift Dt values are listed in Table S6. “Labair+propanol/isopropanol” indicates the standard spikes into the background lab air.

**Fig 3.**
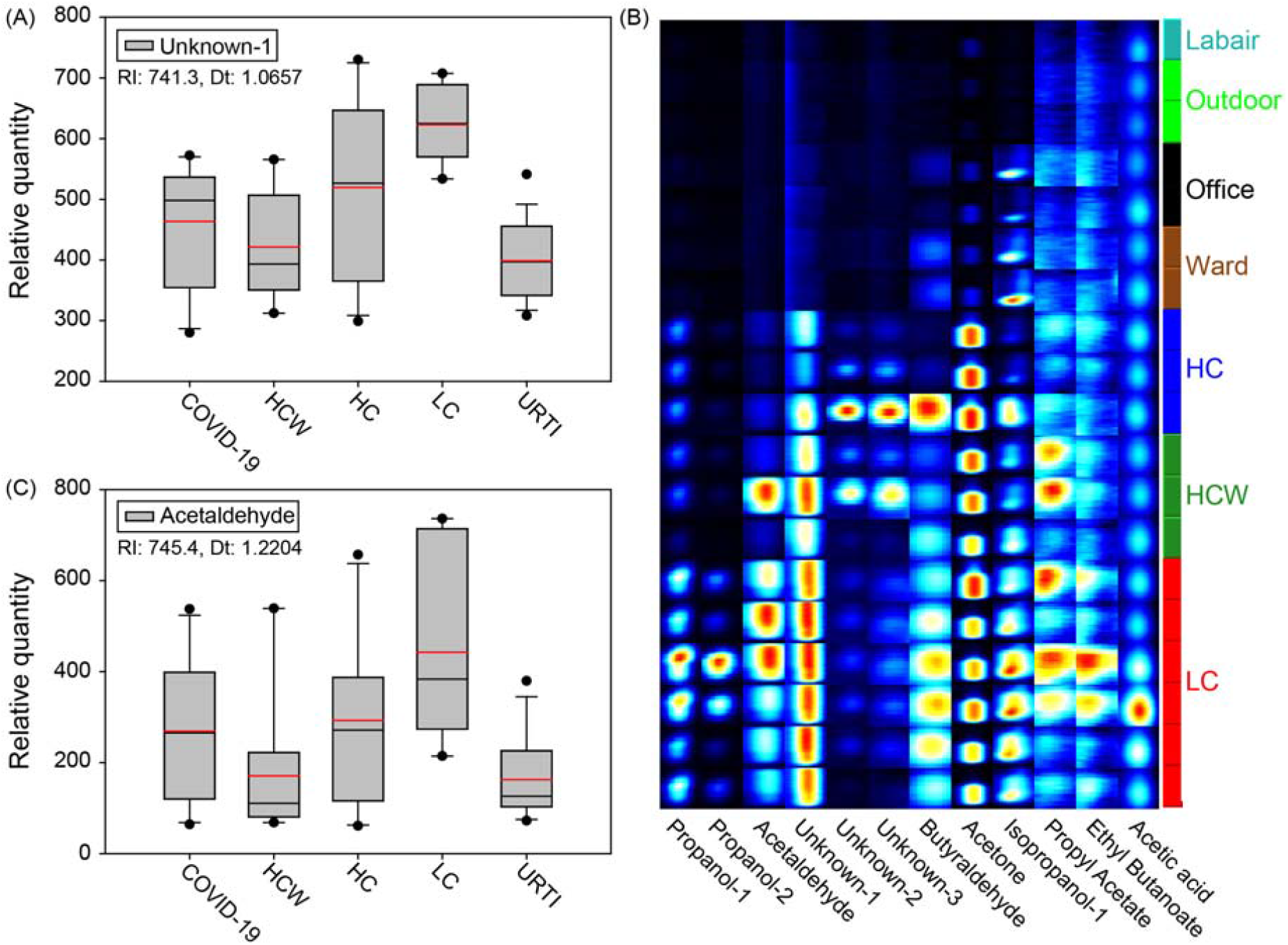
Characteristic breath-borne VOC boxplot profiles for LC patients compared to other groups of subjects: A) Unknown-1 (RI:741.3; Dt: 1.0657) levels of LC patients compared to those in other different groups; B) Acetaldehyde levels of LC patients compared to those in other groups; C) Examples of VOC abundance prints for selected LC patients, controls, various environments. For unknown VOCs, their RI and Dt values are listed in Table S6.

Rapid diagnosis of COVID-19 plays a vital role in controlling the pandemic. Breath-borne VOCs have been investigated as biomarkers for lung cancer^5^, oxidative stress^6^, and many other diseases.^7^ The VOC emission is described as a direct or indirect cause of reactive oxygen species (ROS) present around the cells.^5,7^ When exposed to substances, the rats were observed to shortly emit distinctive VOC profiles in a recent work.^8^ Likewise, when people are infected by SARS-CoV-2, characteristic protein and metabolite changes were observed; and more than 100 lipids were reported to be down-regulated in their blood sera.^9^ Here we show that breath-borne ethyl butyrate (C6H12O2) seems to be characteristic of COVID-19 and URTI (SARS-CoV-2 negative) patients (both of them represent respiratory infections) when compared with subjects with LC, HC, and HCW (Table 1). In contrast, butyraldehyde(C4H8O) becomes a unique biomarker for screening URTI patients (Fig 2; Table 1), as it was observed to be significantly higher in exhaled breath of URTI than those of COVID-19, HC, HCW, LC subjects and also background air samples (p-values<0.001; Table 1). As for screening, a VOC profile of lower level of butyraldehyde but higher level of ethyl butanoate, as revealed here, corresponded to a COVID-19 infection. Higher levels of both butyraldehyde and ethyl butanoate likely result from infections by other pathogens, but not by SARS-CoV-2. For lung cancer, it seems that no such candidate biomarkers were found, although Unknown-1 (RI:741.3; Dt: 1.0657) and acetaldehyde (Table S6) demonstrated the potential when only compared to HCW (p-values<0.008). Nonetheless, it is difficult to eliminate the influences on these emitted VOCs from the individual variations, diet, and medication.^5,7^

Besides the main symptoms such as fever and cough (>80%) as described ^10^, “silent hypoxia”, a condition of oxygen being deprived, was also reported for COVID-19 patients.^11^ Here, we observed a decline of breath-borne acetone(C3H6O) level, and an increase in ethyl butanoate, which could be due to the changes of metabolites resulting from SARS-CoV-2 infections as reported.^9^ The elevated ethyl butanoate level from COVID-19 was possibly transformed from the butyraldehyde, which is easy to be oxidized into butyric acid by oxygen. The butyraldehyde level was shown to be significantly higher for URTI patients. These changes can in turn possibly influence the oxygen demands, which can eventually lead to the “silent hypoxia” problem mentioned. Such a metabolism abnormality resulting from the SARS-CoV-2 infection is likely contributing to the cytokine storms in which levels of protein biomarkers substantially increased, e.g., interleukin (IL)-2, IL-7, macrophage inflammatory protein 1-α, tumour necrosis factor-α, C-reactive protein (CRP) and interleukin-6 (IL-6).^9,12,13,14^ Average breath-borne isopropanol-1 from COVID-19 patients was significantly lower than healthy controls (p-value=0.0329, Table 1). In contrast, higher level of isopropanol-1, other than its isomer isopropanol-2, was detected for URTI patients than the controls (p-value=0.002, Table 1). Isopropanol-1, emitted also by healthy controls, was shown to inactivate SARS-CoV-2 by at least 5.9 Log10 within 30 s at 75% (wt/wt).^15^ Thus, isopropanol-1 produced by the COVID-19 patients, which however varied greatly (up to ∼two orders of magnitude), was used as a self-defense to inactivate SARS-CoV-2 or other pathogens. Whether breath-borne SARS-CoV-2 can be efficiently inactivated depended on the level of isopropanol-1 emitted from a particular patient. Accordingly, the isopropanol-1 level might to some extent affect the viability of SARS-CoV-2 emitted by COVID-19 patients. COVID-19 patients with low levels of isopropanol-1 might serve as a super-spreader of viable SARS-CoV-2. Previously, we have shown that COVID-19 patients emitted millions of SARS-CoV-2 per hour during the earlier stages of the disease.^4,6^ Accordingly, some of those emitted SARS-CoV-2 could have been inactivated by breath-borne isopropanol-1. For rapidly screening COVID-19, machine learning, as used in a previous work^9^, can be further employed to analyzing breath-borne VOC profiles from a larger size of COVID-19 patients in forthcoming research efforts. The findings here could lend considerable help in rapidly screening COVID-19 patients even in their earlier stages without symptoms, given the high sensitivity (1-5 ppb for aldehydes, ketones, and lipids) of GC-IMS. The candidate biomarkers such as ethyl butanoate and isopropanol-1 can be also used to monitor the presence of COVID-19 patients in particular environments, thus serving as an alert for COVID-19. The larger sample size of COVID-19 patients was desired, but impacted by both biosafety concerns and lab constraints due to the highly infectious nature of SARS-CoV-2. This work pioneers a technological advance for rapidly screening COVID-19 using breath-borne VOCs, while shedding new lights on how a COVID-19 patient actively defends against the virus by emitting isopropanol.

## Materials and Methods

### Exhaled breath VOC collection from subjects

A total of 60 subjects were recruited, including 10 COVID-19 patients, 22 non-COVID-19 patients (upper respiratory tract infection, URTI), who had respiratory infections (including pneumonia); 9 healthy health care workers (HCWs) who performed breath sample collection and conducted epidemical tracing with COVID-19 patients; 7 hospitalized lung cancer (LC) patients; and 12 healthy controls (HC) who did not have direct contacts with COVID-19 patients or their associated environments (Table S1-S5). To sample exhaled breath volatile organic compounds (VOCs), 1-L ALTEF gas sample bags (Jensen Scientific Products, Inc., FL, USA) were used. The subjects were asked to exhale into the bag for 2-3 min. For every single expiration, the first half breath was exhaled outside of the bag; and only the remaining breath was collected into the bag, thus ensuring an end-tidal breath collection only from lower parts of the lung. The subjects were asked in advance not to eat, or drink, if needed, only water in 2 hours before the sample collection; and advised to gargle with water right before providing breath samples to avoid the possible interferences from food and mouth sources. Background air samples in subject residing environments for COVID-19 wards, HCW offices, outdoor air for healthy controls, the fever clinics for URTI patients and LC wards for LC patients were collected for quality controls using a gas sampling pump (Dalian Hede Tech Ltd., Liaoning, China). The recruitment of COVID-19 patient was based on a nucleic acid test for SARS-CoV-2 and clinical symptoms provided by a hospital in Beijing. The lung cancer patients were diagnosed by another hospital in Beijing. The URTI patients had clinical flu symptoms, but not SARS-CoV-2 infection as tested by two hospitals in Beijing. Recruitments and further analysis inclusion of both HCW and HC subjects in relation to COVID-19 or URTI were based on no onset clinical symptoms or no lung problems for at least one-month observation time since the sample collection. The ethics involving non-invasive collection of exhaled breath from human subjects was waived due to the urgency of the COVID-19 outbreak investigation; and approved by the Ethics Committee of the Center for Disease Control and Prevention of Chaoyang District of Beijing.

### Analysis of breath-borne VOCs

Breath-borne volatile organic compounds (VOCs) (N=40 species) profiles from these subjects (N=60) and corresponding background air samples (N=30) were analyzed within 12 hours after collection using BreathSpec GC-IMS (G.A.S., Dortmund, Germany) consisting of a gas chromatograph (GC) and an ion mobility spectrometer (IMS). In this study, the GC-IMS was equipped with a Wax column (Restek, Co., PA, USA). The carrier and buffer gas for GC-IMS was nitrogen that was generated by N_2_-Generator (G.A.S., Dortmund, Germany) with a 99.999% purity. For every breath or air sample, the analysis was repeated twice with a measurement variation of 0.1-5.0% by the BreathSpec. The VOCs peak areas were selected and identified using VOCal software with GC-IMS Library (v 0.1.1, G.A.S., Dortmund, Germany) based on their retention index (RI) and drift time (Dt) values. The relative quantities of VOCs (N=40 species) were determined by the peak volumes using the software. The standard liquid sample of propanol (>99.5% purity grade, Concord Tech Co., Ltd, Tianjian, China), isopropanol (>99.7% purity grade, TongGuang Fine Chemicals Co., Ltd, Beijing, China) and ethyl butanoate (99%, Adamas Reagent Co., Ltd, Shanghai, China) after serial dilution were used to confirm and calibrate the VOCs species identification. For a particular sample, the analysis can be finished within 10 min.

## Statistical analysis

As for the VOCs species or peaks, the relative levels of breath-borne VOCs were determined by subtracting the breath-borne values measured from the subjects by those from their associated air environments. The statistical differences in specific relative VOC concentrations between each pair of different subject groups were analyzed via t-test (data exhibited a normal distribution with equal variance) or Mann-Whitney Rank Sum Test (data did not follow a normal distribution or equal variance test failed). The differences of VOCs levels between breath samples and their corresponding air samples were analyzed via paired t-test (data exhibited a normal distribution) or Wilcoxon Signed Rank Test (data did not follow a normal distribution). All statistical tests were performed with Sigmaplot 12.5 (Systat Software, Inc., IL, USA). The average relative levels of breath-borne VOCs, such as acetone, butyraldehyde, isopropanol, ethyl butanoate, and propanol, were 1.86-9.54 times those measured from the background air. A p-value of less than 0.05 indicated a statistically significant difference at a confidence level of 95%.

## Data Availability

All data are available in the main text or the supplementary materials.

## Supporting Files

**Fig. S1.**
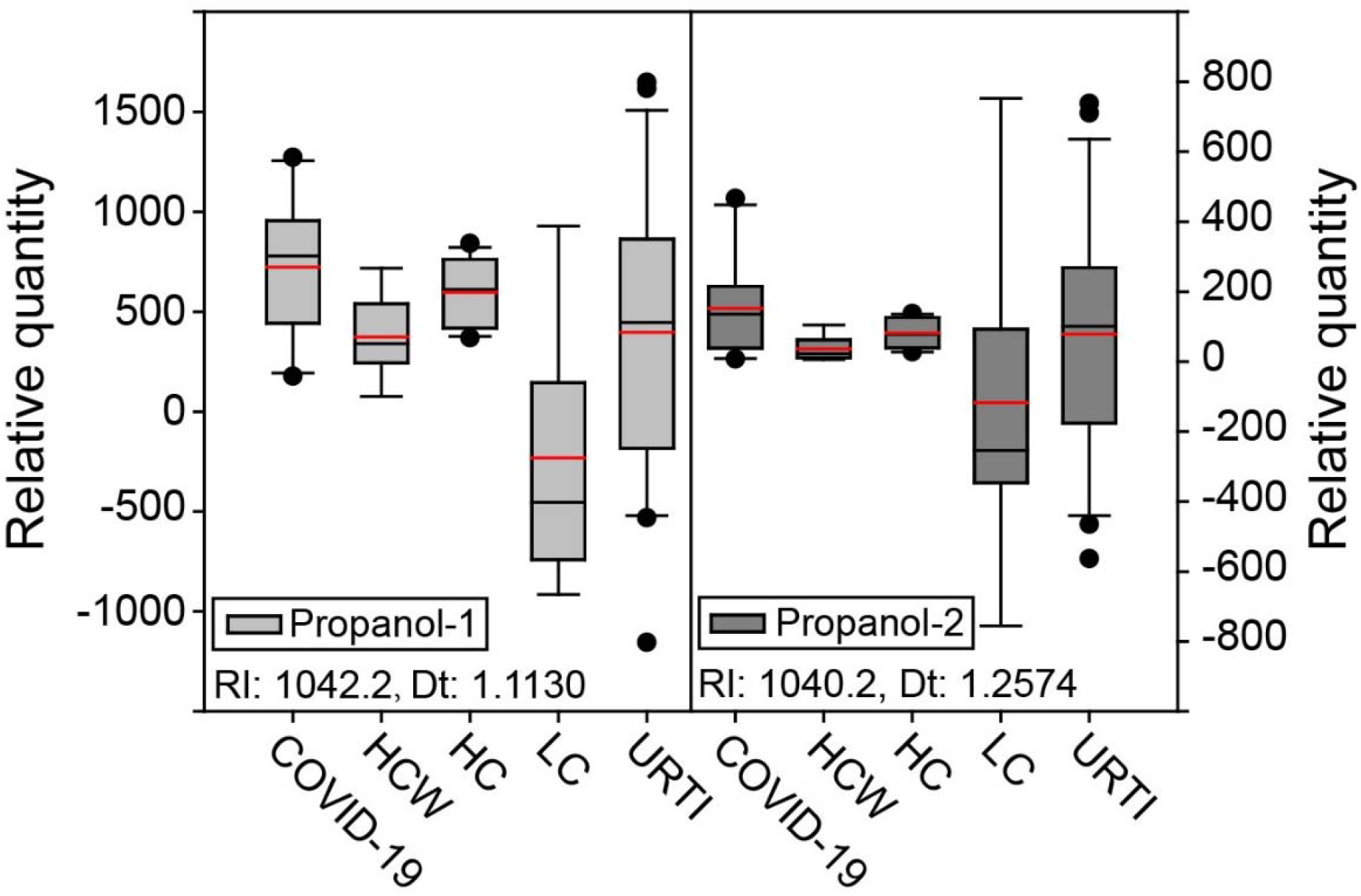
Relative levels of breath-borne propanol-1 (RI: 1042.2; Dt: 1.1130) and propanol-2 (RI: 1040.2; Dt: 1.2574) measured from COVID-19 patients (N=10) compared to HCW (N=9), URTI (N=22), LC(N=7), and HC (N=12). All VOC levels were subtracted by the corresponding background ones.

## Data S1. (separate file)

The excel file contains Table S1 (Medical records of COVID-19 patients and their bio-information), Table S2 (Medical records of upper respiratory tract infection (URTI) subjects and their bio-information), Table S3 (Bio-information of Health Care Workers (HCW)), and Table S4 (Medical records of lung cancer (LC) patients and their bio-information), Table S5 (Bio-information of Healthy Control (HC) subjects), Table S6 (The relative levels of breath-borne VOCs measured from COVID-19, LC, HCW, HC, and their associated backgrounds), Table S7 (The 95% CIs for relative levels of breath-borne VOCs measured from five groups of subjects and their associated background air samples).

## Author contributions

M.Y., and J.M. contributed to the study design. H.C., X.Q., J.M. contributed to sample collection and experiments. H.C. performed all VOC analyses. J.M., C.Z., H.F. contributed to patients’ recruitment, and clinical management. H.C., M.Y., J.M., X.Q., C.Z. contributed to data analysis, data interpretation, figure preparation and literature search. M.Y. wrote the manuscript draft, and all authors revised the manuscript. All authors reviewed and approved the final version of the report.;

## Data and materials availability

All data are available in the main text or the supplementary materials.

